# Variants in SARS-CoV-2 Associated with Mild or Severe Outcome

**DOI:** 10.1101/2020.12.01.20242149

**Authors:** Jameson D. Voss, Martin Skarzynski, Erin M. McAuley, Ezekiel J. Maier, Thomas Gibbons, Anthony C. Fries, Richard R. Chapleau

## Abstract

**Introduction:** The coronavirus disease 2019 (COVID-19) pandemic is a global public health emergency causing a disparate burden of death and disability around the world. The molecular characteristics of the virus that predict better or worse outcome are largely still being discovered.

**Methods:** We downloaded 155,958 severe acute respiratory syndrome coronavirus 2 (SARS-CoV-2) genomes from GISAID and evaluated whether variants improved prediction of reported severity beyond age and region. We also evaluated specific variants to determine the magnitude of association with severity and the frequency of these variants among the genomes.

**Results:** Logistic regression models that included viral genomic variants outperformed other models (AUC=0.91 as compared with 0.68 for age and gender alone; p<0.001). Among individual variants, we found 17 single nucleotide variants in SARS-CoV-2 have more than two-fold greater odds of being associated with higher severity and 67 variants associated with ≤ 0.5 times the odds of severity. The median frequency of associated variants was 0.15% (interquartile range 0.09%-0.45%). Altogether 85% of genomes had at least one variant associated with patient outcome.

**Conclusion:** Numerous SARS-CoV-2 variants have two-fold or greater association with odds of mild or severe outcome and collectively, these variants are common. In addition to comprehensive mitigation efforts, public health measures should be prioritized to control the more severe manifestations of COVID-19 and the transmission chains linked to these severe cases.

## Introduction

Since the coronavirus disease 2019 (COVID-19) pandemic emerged, humans have faced unprecedented disruption from the newfound obligate parasite. Within the United States alone, the unwelcome guest has already caused an estimated 2.5 million years of life lost.(1) Beyond the United States, the global burden is substantial and growing, but it is not uniform; continents, nations, communities, families, and patients are all affected differently. Understanding the basis for this variability is an important global health priority.

One of the most common measures to describe the severity of COVID-19 is the infection fatality ratio (IFR) or the number of deaths for every infection. There are widespread differences in IFR between studies,(2-4) and this heterogeneity is not due to chance alone (p<0.001).(2) One meta-analysis of 26 studies estimated an IFR of 0.68% (0.53-0.82%) while cautioning this was likely an “underestimate” and another meta-analysis with 61 studies estimated a median IFR of 0.26%, outside the range of the first meta-analysis.(2, 3)

Other studies have suggested the variability in infection fatality ratio is more consistent when infections are stratified by age, but is more likely to vary between studies when considering the population above age 65.(4-6) In fact, one paper even estimated that “differences in age structure of the population and the age-specific prevalence of COVID-19 explain nearly 90% of the geographical variation in population IFR.”(4)

Although vulnerable age groups should be protected as recommended by the U.S. Centers for Disease Control and Prevention (CDC), there are likely more factors at play in the IFR than age alone. First, there is variability among some younger groups. In the most recent meta-analysis among those age <70, IFR estimates varied from 0.00% to 0.31% with a median of 0.05%.(3) Second, even within the same hospital systems, there are strong time trends in case fatality rate after adjusting for baseline characteristics/risk (7) even at the national scale (8), which authors attributed to better treatment. Unless baseline risk adjustments were inadequate, there are likely time trends in IFR, which would also help reconcile the disparate estimates noted in the two meta-analyses cited above.

Another possibility is severe acute respiratory syndrome coronavirus 2 (SARS-CoV-2) virulence could differ across geographic locations because some local strains have higher or lower virulence than others. Previous reports have argued that evolution of attenuated strains is expected for RNA viruses and that “determination of naturally circulating attenuated SARS-CoV-2 variants is an urgent matter.” (9) Previous publications on evolution of virulence observed that pathogens transmitting with a respiratory route (unlike vector-borne diseases) typically evolve toward lower virulence after emergence because healthier hosts tend to engage in more social contact.(10)

Using publicly available data from GISAID (Global Initiative on Sharing Avian Influenza Data), we interrogate the relationship between SARS-CoV-2 variants and associated patient outcomes in the GISAID metadata. We differentiate between severe and mild patient outcomes and utilize a logistic regression model in order to better understand how viral genomic SARS-CoV-2 variants are linked with COVID-19 patient outcomes.

## Methods

### Variant Alignment and Variant Calling

SARS-CoV-2 genome sequences were obtained from GISAID (Global Initiative on Sharing All Influenza Data) on October 21, 2020 (11, 12), (GISAID Acknowledgments Table, Supplementary Table 1). GISAID sequences were filtered to include those of human origin. FASTA sequences were aligned to the reference sequence, Wuhan-Hu-1 (NCBI: NC_045512.2; GISAID: EPI_ISL_402125) using Minimap2 (version 2.17).(13) Resulting VCF (Variant Call Format) files were annotated using SnpEff (version 5.0) and filtered using SnpSift.(14, 15) The shell scripts used for variant alignment and variant calling, along with the Python scripts used to perform the steps described below, are available on GitHub at https://github.com/mskar/variants.

### Metadata Preprocessing and Cohort Building

Raw GISAID patient data was parsed from the JSON file using Python (version 3.8.2). Patient outcomes were then aggregated into positive (“Mild”) outcomes or negative (“Severe”) outcomes (detailed in Supplemental Figure 1). Briefly, “Mild” outcomes included: Outpatient, Asymptomatic, Mild, Home/Isolated/Quarantined, and Not Hospitalized. “Severe” outcomes included: Hospitalized (including severe, moderate, and stable) and Deceased (Death). Patient outcomes that were unclear or empty were not included in our analyses.

**Figure 1:**
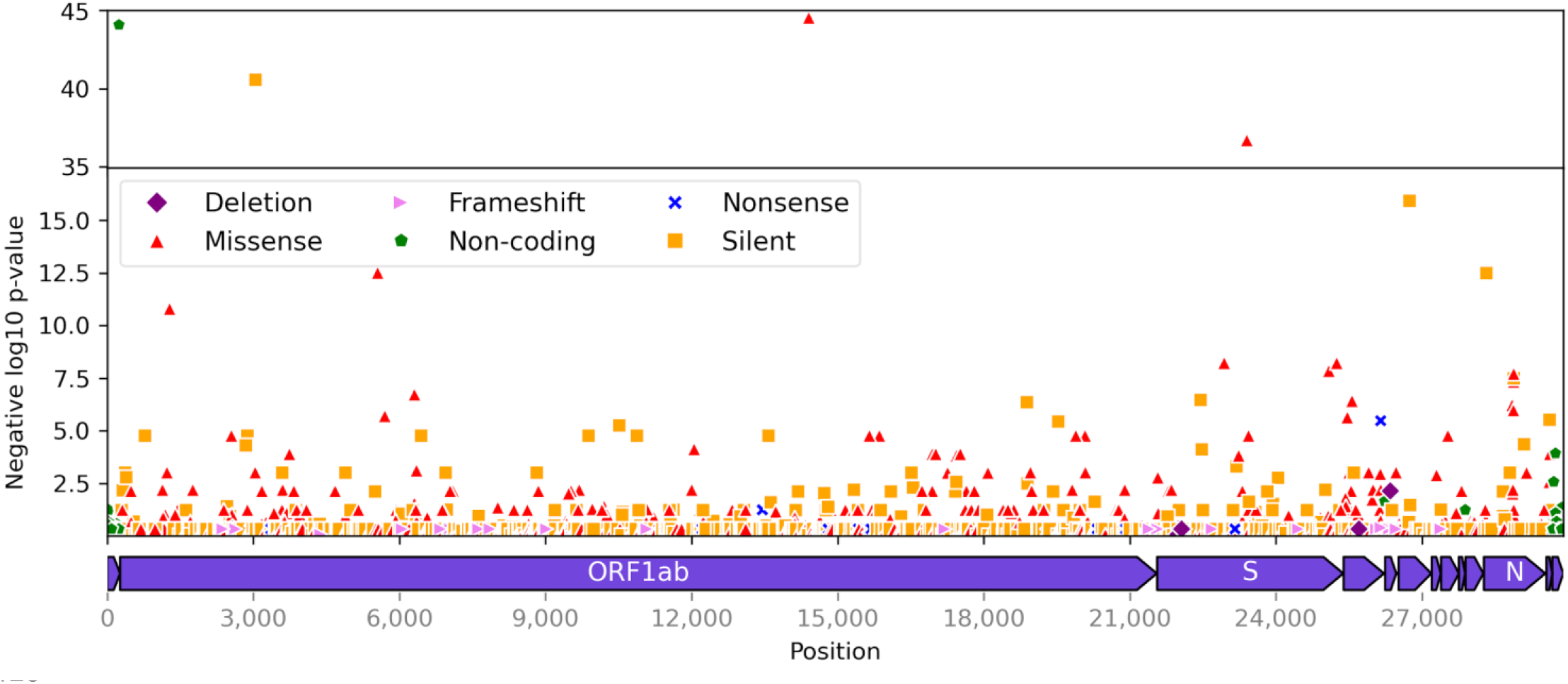
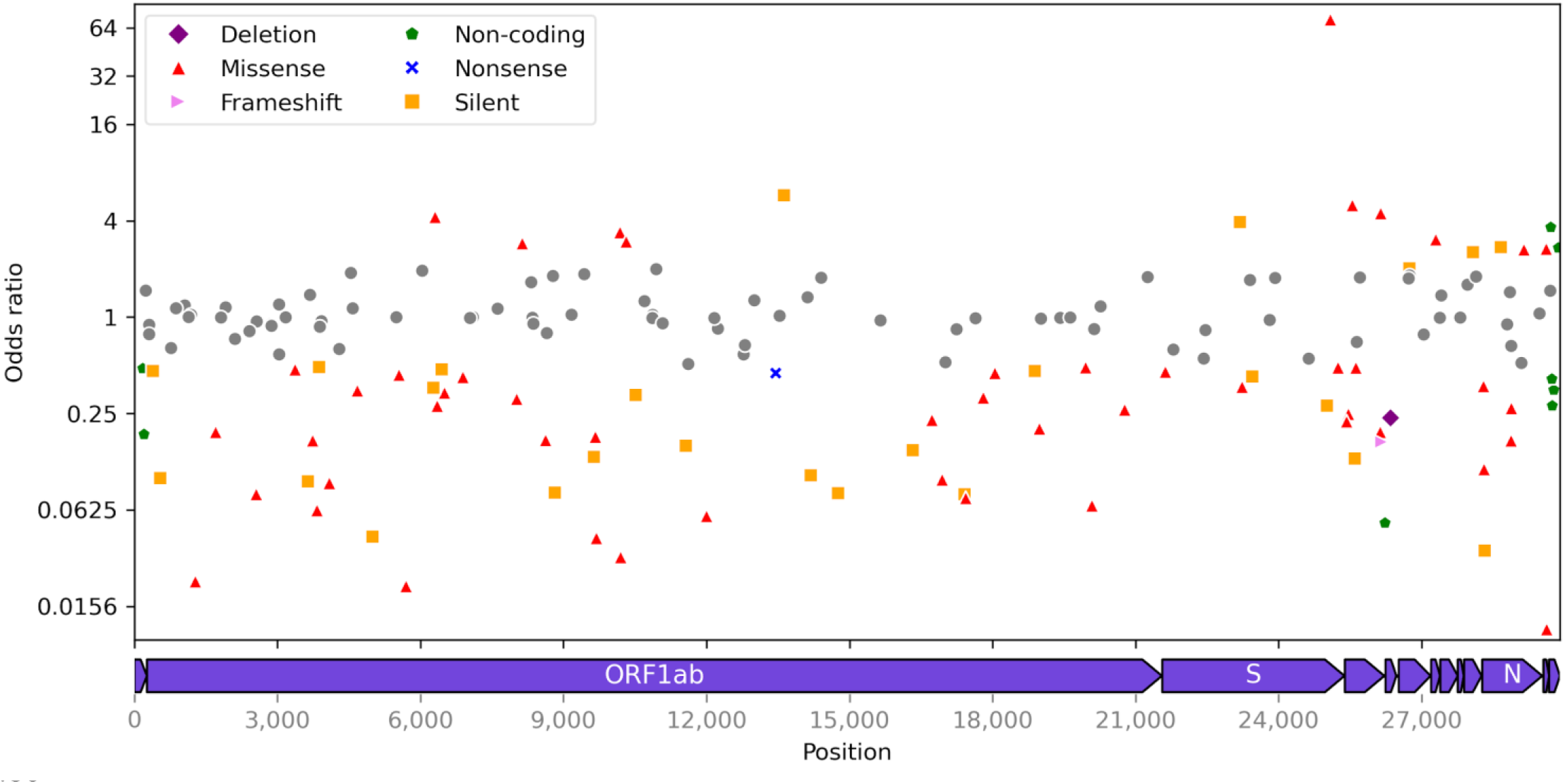
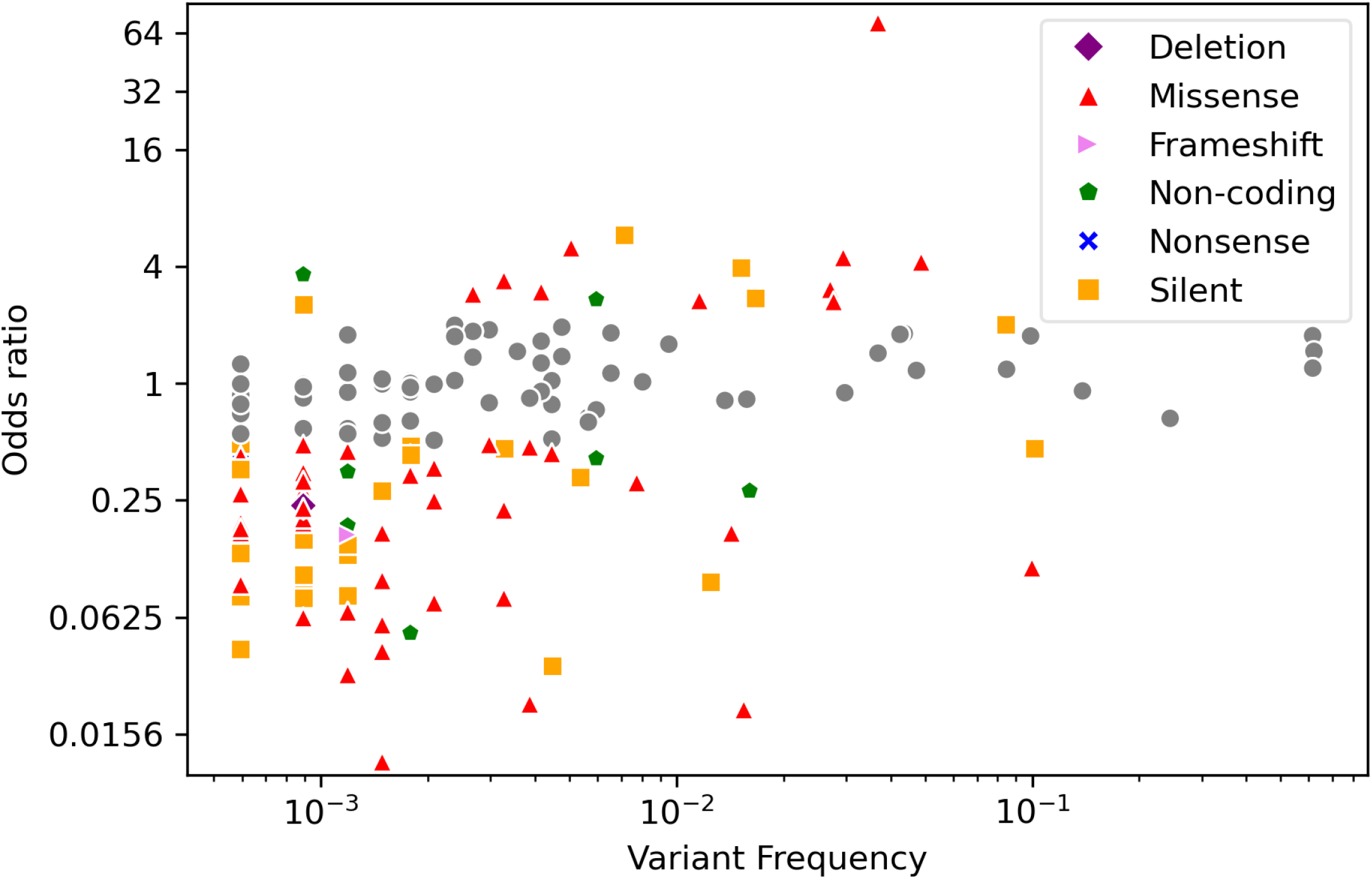
Overview of SARS-CoV-2 variants selected from GISAID data (n = 1595168). A) Negative log10 p-values of variant association (chi-square test) with “Severe” outcome group (hospitalized, deceased, etc.) plotted against position of variants (n = 4484) in the SARS-CoV-2 genome. B) Odds ratios (log2 scale) of “Severe” versus “Mild” (outpatient, asymptomatic, etc.) outcome groups plotted against the positions of variants with odds ratios not equal to one (n = 169) in the SARS-CoV-2 genome. C) Odds ratios (log2 scale) of “Severe” versus “Mild” outcome groups plotted against log10 frequency of variants (n = 169) in the patient subpopulation (n = 3363) without missing variables. Points are labeled by mutation type (red: missense, green: non-coding, orange: silent, yellow: nonsense, purple: deletion).

### Variant and Metadata Modeling

Annotated VCF files were parsed, pivoted to wide format, and joined with GISAID patient data using Pandas (version 1.0.3).(16) Logistic regression models with the default L1 penalty (Lasso regularization) were fit to the patient (rows) and variant (columns) data using Scikit-learn (version 0.23.2).(17) Logistic regression model Area Under the Curve (AUC) and accuracy values were calculated using Scikit-learn.(17) Models were persisted as pickle files using joblib (version 0.14.1).

### Plotting and Statistical Analysis

Scatter and bar plots were created using Pandas (version 1.0.3),(16) Matplotlib (version 3.2.1),(18) and Seaborn (version 0.10.1).(19) Logistic regression model AUC p-values and Chi-square test p-values for association of variants with “Severe” outcomes were obtained using Scipy (version 1.5.0).(20) Variant frequency was calculated using Pandas.(16) Genome position tracks were added to scatterplots using DNA Features Viewer (version 3.0.3).(21) ROC curves were plotted using Scikit-learn (version 0.23.2),(17) and Matplotlib.(18) Logistic regression model Area Under the Curve (AUC) and accuracy values were calculated using Scikit-learn.(17)

## Results

### Sample population characteristics

We collected a total of 155,958 viral genomes along with clinical metadata. The metadata included numerous entries whereby the severity of the condition could not readily be discerned. For example, “recovering”, “recovered and released”, and “mild symptoms inpatient for observation” were found in the raw data and were not included. The full downloaded dataset included 148,121 entries with empty or unknown clinical observations, and 4,200 entries for which clinical severity could not be classified. From the remaining 3,637 sequences with clear severity indications, we generated two classes (Supplemental Figure S1) by recoding the observational metadata into consistent terminology and creating a “Severe” class of “deceased”, “hospitalized”, “ICU”, and “pneumonia” (n=2,870); and a “Mild” class of “outpatient”, “mild”, “epidemiology study”, “asymptomatic”, “screening”, and “stable in quarantine” (n=767). 85% of these genomes had at least one variant associated with patient outcome. Viral sequences were obtained from the six major geographical regions in GISAID between January and October 2020 (Supplemental Figure S2).

### SARS-CoV-2 variants associated with “Severe” / “Mild” outcome categories

The overwhelming majority of variants in the SARS-CoV-2 genomes assessed were rare, with only 12 common variants with at least a 5% minor allele frequency (Figure 1C). Two of these common variants (C26735T and C28311T) were associated with “Severe” or “Mild” outcomes, as measured by having an odds ratio of greater than 2 or ≤ 0.5, respectively. We also observed 84 of 157 rare variant associations with “Severe” or “Mild” outcomes. Collectively, 17 variants were associated with “Severe” classification with at least an odds ratio of 2, while we found 67 associations with “Mild” classification (OR≤ 0.5). The variants associated with outcomes were distributed across the genome, including the strongest “Severe” association within the C-terminal end of the spike protein (Figure 1B). The majority of variants characterized here were transitions (121, 71%), with 47% of those transitions (79) being C>T (Supplemental Figure S3). All individual variant associations and frequencies are reported in Supplementary Table 2.

### Predicting clinical outcomes for patients based upon clinical metadata and viral genomics

Age and gender have been previously reported to be predictive of clinical outcomes.(22) Our observations predicting “Severe” outcomes confirm these prior associations (Figure 2); however we found that the c-statistic for predictions based on age or age and gender are only slightly better than random chance at 0.677 (0.642-0.712) and 0.679 (0.644-0.714), respectively. We hypothesized that viral genomic variants in the virus could also contribute to severity classification. When accounting for the region of collection, we observed an increased c-statistic to 0.817 (0.817-0.818). Previous observations show that regions tend to be dominated by individual viral clades, and we found that adding clade to an age/gender/region model resulted in moderate improvement of accuracy (81% vs. 86%, respectively) with a nearly identical AUC of 0.818 (0.817-0.818). While the difference between region and clade appears insignificant, adding clade-level information increased the predictive ability of our model beyond age and gender alone. We then considered whether variant-level information would further improve model performance. We found that substituting clade with 4,499 genomic variants in the model increased the c-statistic to 0.911 (0.910-0.911), significantly improving predictability for clinical outcomes. In addition to the improvement in c-statistic, we compared the accuracy of our predictions, which started at 81% for the age-only model and improved to 86% and 88% for each additional step in the model building before finally reaching a maximum at 91% accuracy for the age/gender/region/variant model.

**Figure 2:**
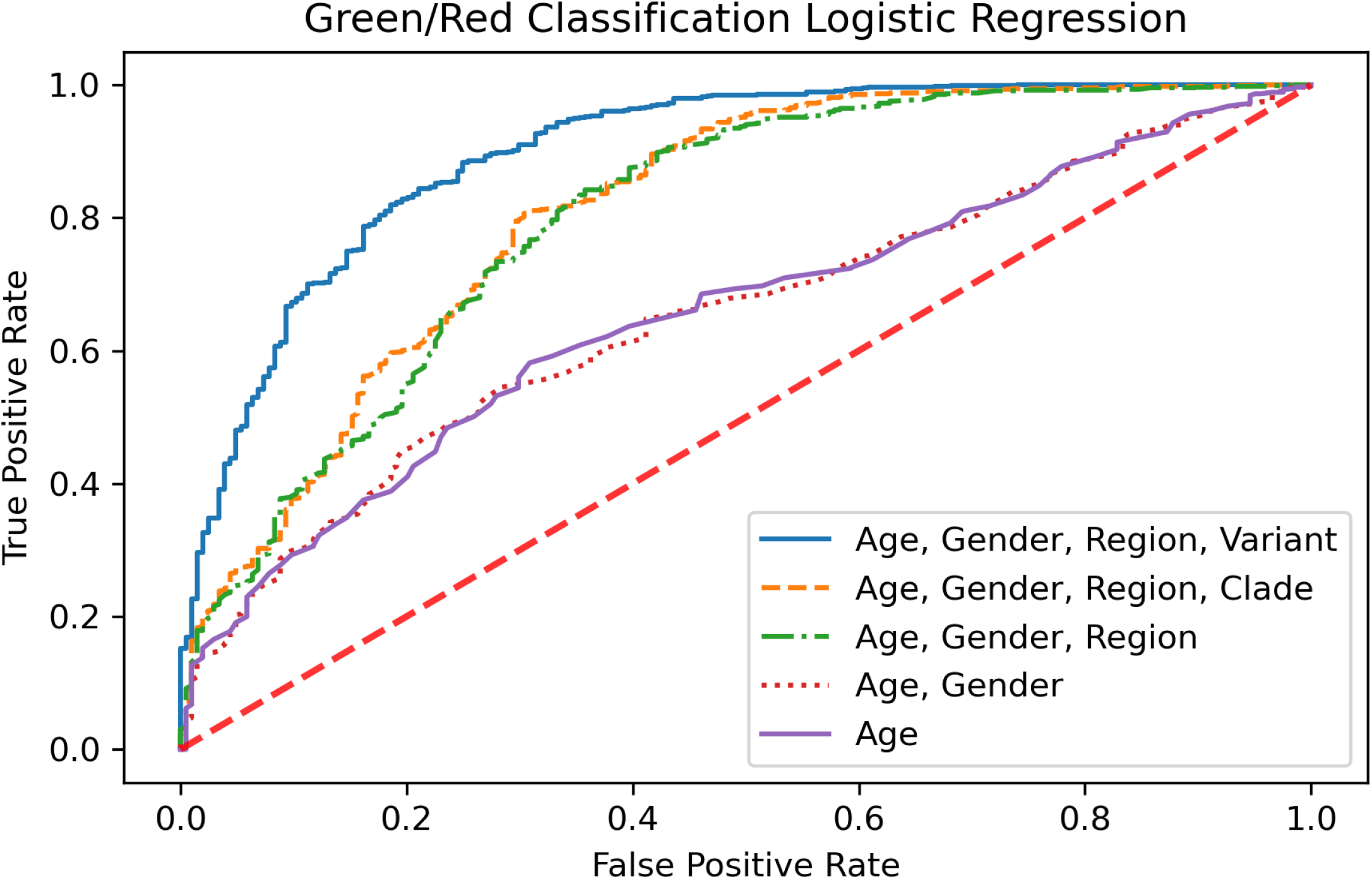
Comparison of nested logistic regression models. Models are labeled based on the predictor variables (purple solid line: age; red dotted line: [age, gender], green dash-dotted line: [age, gender, region], orange dashed line: [age, gender, region, clade], blue solid line: [age, gender, region, variant]) used to predict whether SARS-CoV-2 patients (n = 3363) belong to “Severe” (hospitalized, deceased) or “Mild” (outpatient, asymptomatic, etc.) outcome groups.

Classifications based solely on age or on age and gender resulted in insignificant odds ratios in our models. Both models had the same odds ratio (4.4), confidence interval (2.8-6.0), and p-value (0.072). However, the other three models all had significant odds ratios with p-values less than 0.0001. Consistent with the AUC results, the odds ratio was greatest for the full model (age/gender/region/variant) followed by age/gender/region/clade and finally age/gender/region (odds ratios: 12.3 (11.8-12.8), 8.4 (8.0-8.9), and 8.0 (7.6-8.4), respectively). Similarly, the negative likelihood ratio for the full model displayed a large reduction in the likelihood of a patient classified as “Mild” developing “Severe” symptoms (-LR=0.039) as compared to a moderate reduction in the post-test outcomes for the age-only or age and gender models (-LR=0.231 for both models).

## Discussion

We demonstrate that including genomic viral variants can substantially improve classification of COVID-19 patient outcomes as compared with models using only age and region. Moreover, in our models, we observe that some individual variants are particularly important with substantial associations with severity, and that collectively these variants are not rare.

Associations between viral genomic variants and patient outcomes are not unexpected. Consistent with known patterns in the evolution of virulence in RNA viruses (9, 10, 23), we would expect many common strains have differing association with patient severity by this point in the pandemic by chance alone and even as sampling is more likely to occur with severe outcomes, variants correlated with mild outcomes are still being identified.

Though respiratory pathogens often evolve toward lower virulence, there have been historical exceptions.(24) Modeling future fitness landscapes suggests that even partial isolation of symptomatic cases can substantially reduce deaths with less transmission in the short term. Importantly, this isolation can also potentially alter the evolutionary path by favoring less virulent strains.(24) Alterations in virulence can happen with a small number of selections. For example, *E. faecalis* evolved from a pathogen to a commensal strain in 15 passages in a worm model, but most of the worm phenotype changed after just 5 rounds of bacterial selection.(25) A mouse experiment showed higher virulence after 10 passages of a modified SARS-CoV-2 virus with an additional ∼1% drop in mouse body weight with each selection (26). Based upon these findings and the large number of passages in the human outbreak, it can be expected that significant evolution affecting virulence could occur in the SARS-CoV-2 genome.

Indeed, others have previously found and characterized individual variants with *in vitro* assays (27) or provided correlates of severity with any change in protein coding,(28) or genomic correlates of mortality.(29) We have taken a comprehensive approach to describe all variants associated with mild or severe outcome regardless of whether it is synonymous. There are challenges in identifying signatures of selection in non-coding regions, but “for RNA viruses…critical aspects of the life cycle rely on molecular processes that are not reflected in protein sequence.”(30) Empiric tests of selection and structural modeling of RNA and protein interaction can identify regions under selection without using the ratio of nonsynonymous to synonymous mutations.(30) Studies of selection in SARS-CoV-2 have also advised not to underestimate the role of synonymous substitutions.(31) Beyond RNA molecule interactions, there also appear to be selective pressures on which codon is used for an amino acid, which could be attributed to tRNA abundance (32) or could be related to broader patterns of host RNA editing involving deamination and similar mechanisms. (33-35) Alternatively, a variant correlated with severity might represent an epiphenomenon that is linked to multiple variants that each have a smaller association with severity. Regardless of the applicability of these explanations for why synonymous changes could be important indicators of virulence, we wanted to take an agnostic approach to characterize all variants correlated with severity so that they could be further resolved with additional study and additional surveillance (particularly among asymptomatic cases). These studies, in addition to comprehensive prognostic study, could better clarify how unexpected a patient’s severity was as compared with additional risk factors.

For example, one mutation we identified (C13620T) is associated with 5.9 times the odds of severe disease. Although it does not result in any change in an amino acid of the NSP12 (RNA-dependent RNA polymerase), it could result in altered expression. Because NSP12 is required for the transcription of all viral RNA in coronaviruses (36), increased replication could increase virulence.

Other mutations were nonsynonymous. We identified two previously reported spike mutations, V1176F (G25088T) and S477N (G22992A), as important indicators for COVID-19 disease severity. Recent protein modeling studies have indicated that both mutations cause favorable energetic changes that result in a more flexible Spike protein and can change RBD-ACE2 binding. Importantly, both mutations have been associated with higher mortality rates, and are therefore expected to have significant impacts on public health.(37, 38) Other mutations were observed less commonly, but could still be relevant for understanding higher viral pathogenesis. The G26144T causes amino acid change G251V in Orf3a and was associated with 4.3 times the odds of severe disease. Protein trafficking is a complex multifactorial process (39) and Orf3a functions as a modulator of the trafficking properties of the spike protein of SARS-1 and is dependent on the protein-protein interaction of Orf3a and S.(40) A structural analysis of the G251V in Orf3a of SARS-CoV-2 results in significant changes in the overall protein structure and weaker affinity for both the S and M proteins with Orf3a. One possible outcome of the weaker Orf3a-S and Orf3a-M interaction could be an increased Orf3a-TRAF3 interaction resulting in increased activation of the NLRP3 inflammasome by promoting TRAF3-dependent ubiquitination of ASC.(41, 42) Thus, the altered protein-protein interaction of the G251V Orf3a may impact the trafficking of Orf3a resulting in a higher propensity for inflammatory cytokine activation.

We also identified that the C28311T mutation was associated with a lower odds ratio of 0.14. This mutation lies within the probe of the N1 assay in the CDC’s PCR assay (43), creating a P13L change in the nucleocapsid protein. A previous study evaluated how this mutation may alter protein-protein interaction and proposed it impacted virus stability, potentially contributing to lower pathogenesis.(44) Additional follow-up studies will help to illuminate effects of these variants on viral fitness, infectivity, host response, and evolutionary trajectory.

Identifying genetic variants associated with outcomes could provide mechanistic understanding of the life cycle.(45) Efforts such as the CDC’s annual influenza surveillance rely upon understanding those key genetic variants to predict seasonal intensity and attempt to develop effective countermeasures (vaccines) (https://www.cdc.gov/flu/weekly/overview.htm).

Although deep molecular insights are important, they are not necessary for public health applications. The CDC offers symptom-based criteria for prioritizing testing and symptom-based criteria are also included in recommendations for prioritizing contact tracing (https://www.cdc.gov/coronavirus/2019-ncov/php/contact-tracing/contact-tracing-plan/contact-tracing.html#). By prioritizing cases and contacts with symptoms (as compared with asymptomatic cases and their asymptomatic contacts) in addition to the recommended global mitigation efforts, there could be relative selective pressure against strains that are more likely to cause symptoms. This could favor the emergence of attenuated strains over the long term.

The COVID-19 pandemic demonstrated the limitations of the global healthcare system in intensive care units, mechanical ventilators, and emerging therapeutics and other medical countermeasures.(46, 47) Early in the outbreak, cities such as New York became inundated with infections and their ability to adequately sort and treat patients was quickly overwhelmed.(48) The existence of a rapid and accurate tool that could help identify COVID-19 patients or clusters that are more likely to experience severe symptoms or require intensive medical resources (e.g., inpatient hospitalization and ventilation) may be able to help healthcare systems allocate resources to the regions with the most critical needs. Therefore, by providing a molecular risk factor for more severe outcomes, these findings could help prioritize limited treatment supplies to those at greatest risk, particularly as therapeutic interventions for infectious disease often need to be given early in the disease course (e.g., empiric antivirals for influenza).

There are limitations with our analyses. First, the SARS-CoV-2 genomes uploaded to GISAID are not necessarily representative of all circulating genomes, which can introduce a selection or sampling bias into our analyses based on region, patient severity, or other unmeasured factors. In Supplemental Figure S2 we show the sampling patterns over time by our patient severity categorization. We sought to mitigate these limitations by eliminating the categories that had ambiguous severity (e.g., “live” or “recovered”), and adjusting the associations for known confounders. In addition, we do not seek to make causal claims about any specific viral genomic variant. In aggregate these variants are predictive of outcome and the candidates we identify can be further studied using molecular and other methods.

In summary, we have demonstrated that some SARS-CoV-2 genomic variants are strong predictors of COVID-19 disease severity, and these variants appear to be commonly circulating. This study provides a rationale for prioritizing control efforts for cases and populations manifesting with unusually high severity, consistent with symptom-based criteria for testing used by the CDC. Longitudinal monitoring of genomic variants within a novel pathogen such as SARS-CoV-2 will be important for understanding drivers and effects of its evolution and ultimately, its spread or control.

## Data Availability

Data is available from GISAID. The shell scripts used for variant alignment and variant calling, along with the Python scripts used to perform the steps described in the manuscript should be accessed on GitHub at https://github.com/mskar/variants

https://www.gisaid.org/

https://github.com/mskar/variants

## Acknowledgements

The authors gratefully acknowledge the contributors, originating and submitting laboratories of the sequences from the GISAID EpiCoV Database (Elbe and Buckland-Merrett, 2017; Shu and McCauley, 2017), the basis of this research. A detailed list of contributing labs to GISAID is available in the Supplementary Information.

## Supplemental Figures

**Figure S1:**
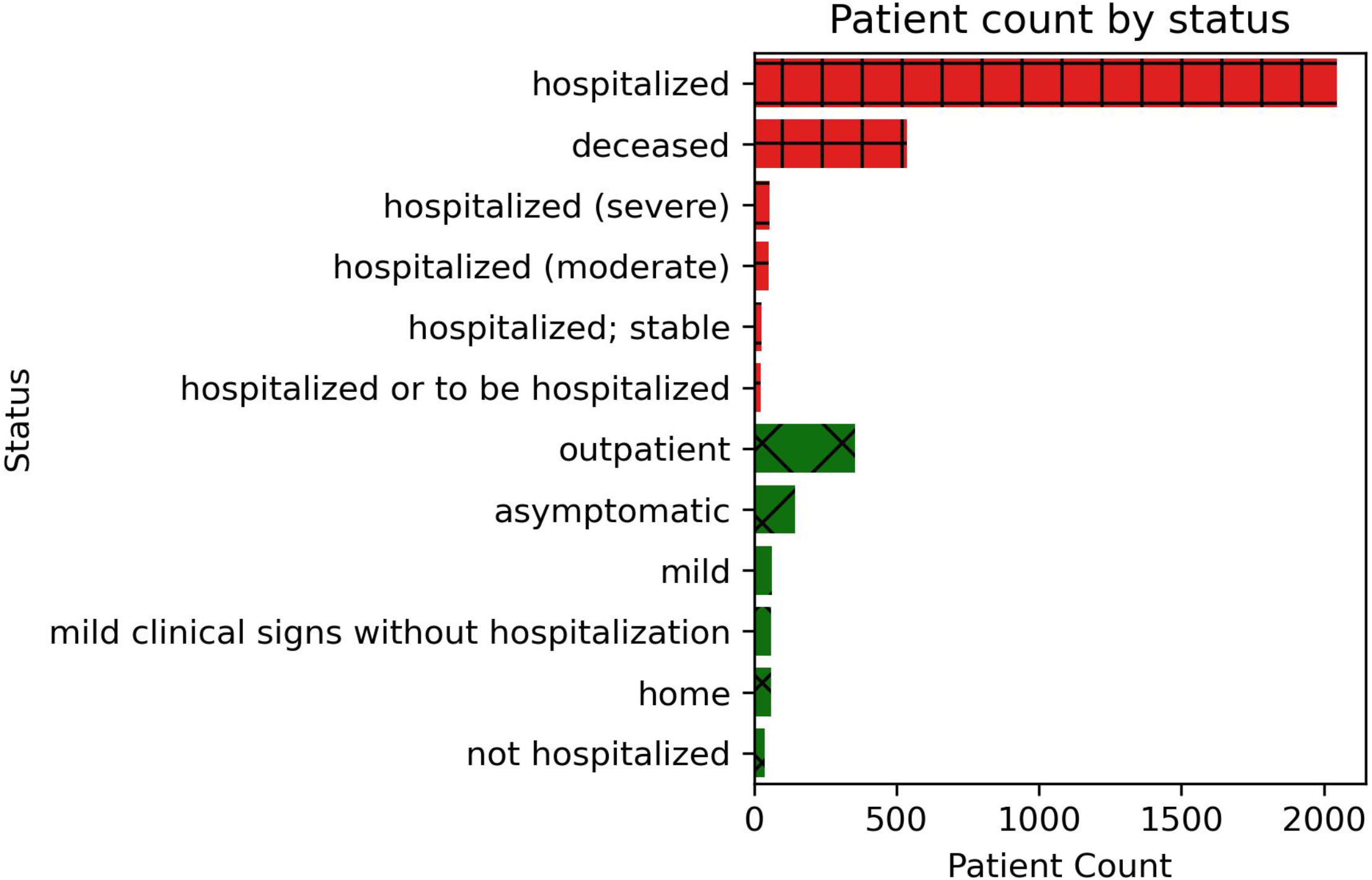
SARS-CoV-2 patient (n = 3611) composition of “Severe” and “Mild” outcome groups. Bars are labeled by group membership (red vertical-horizontal hatch: [severe, symptomatic, deceased], green diagonal hatch: [live, released, mild, recovered, asymptomatic]).

**Figure S2:**
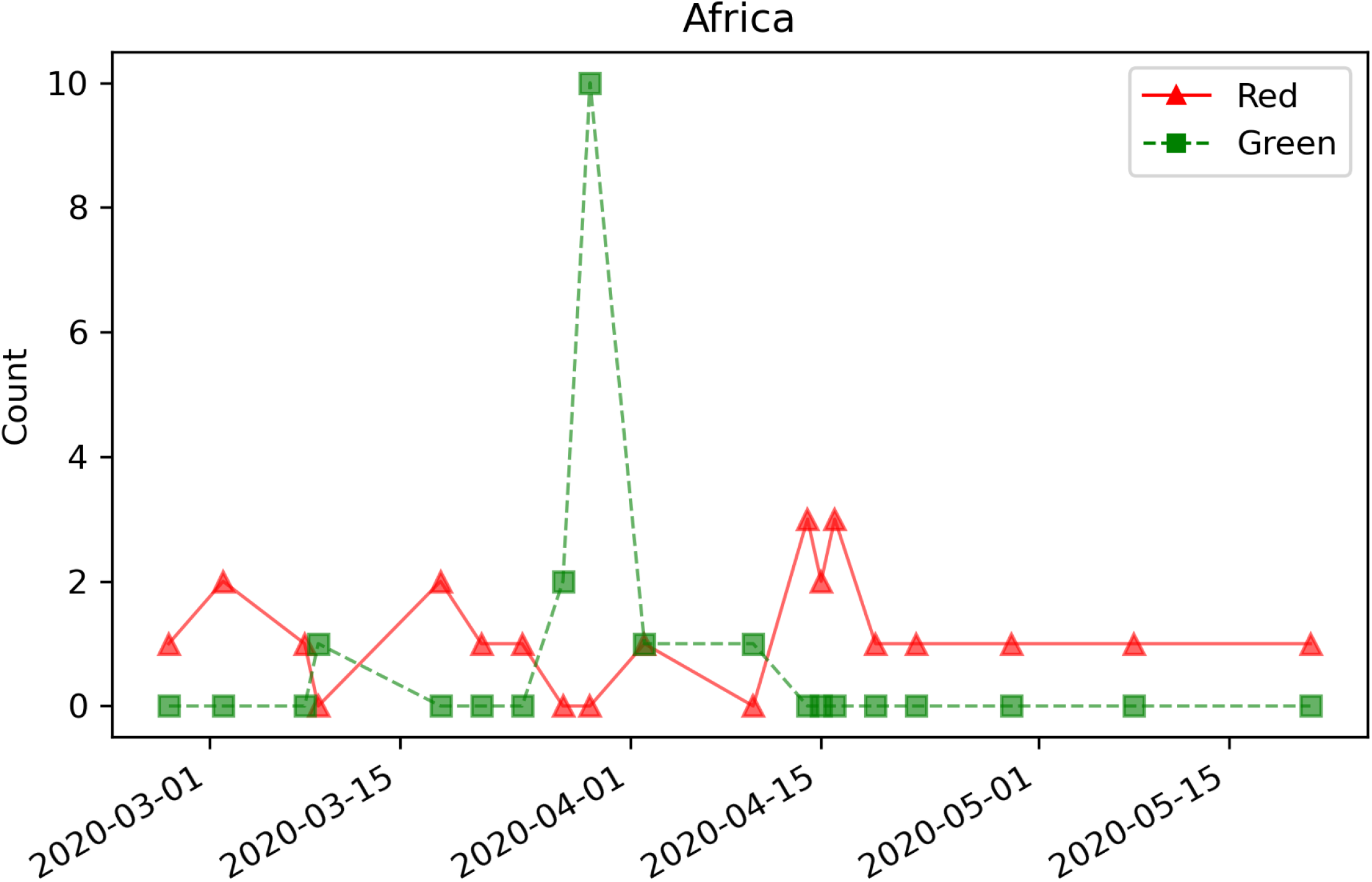
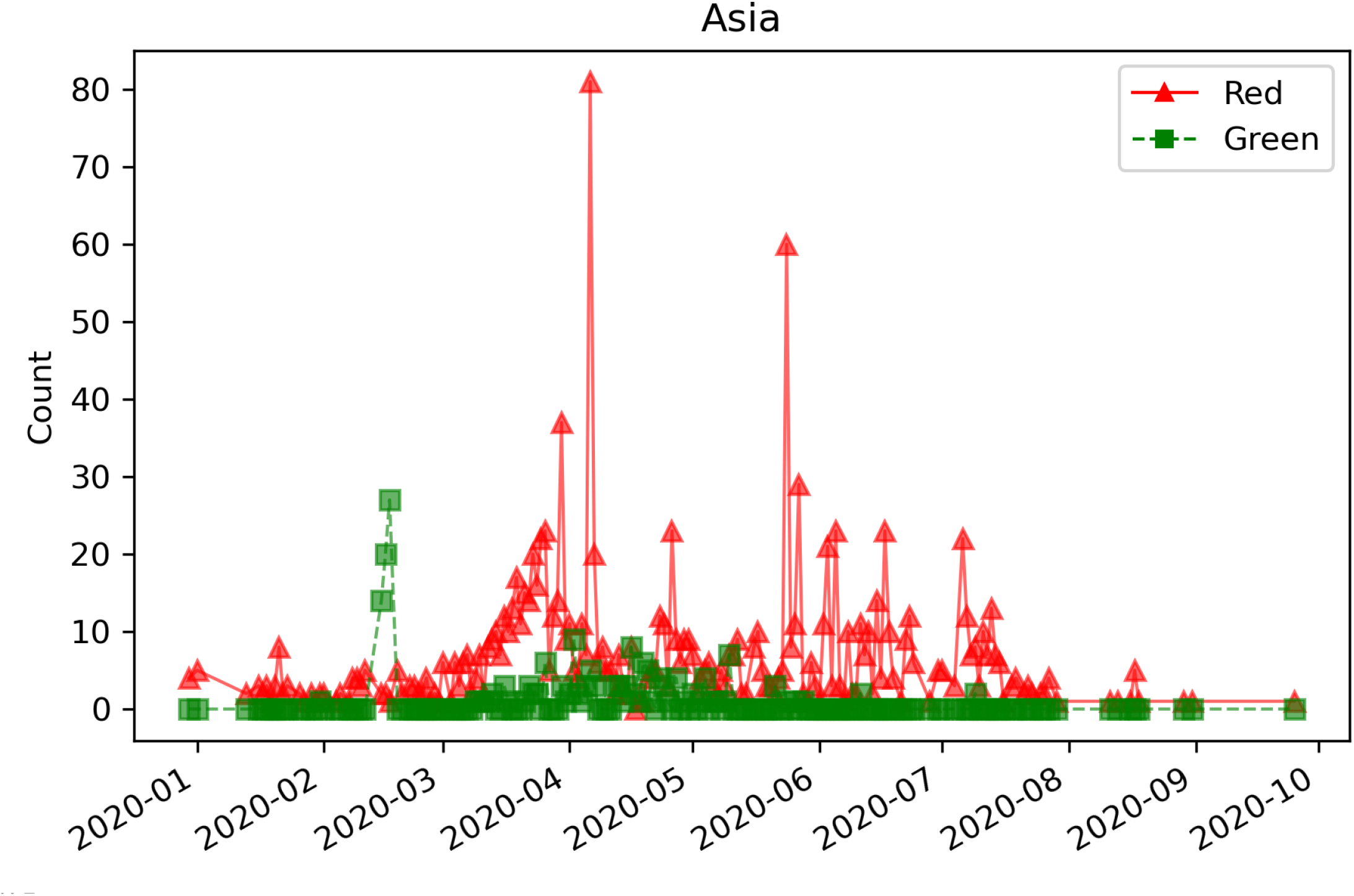
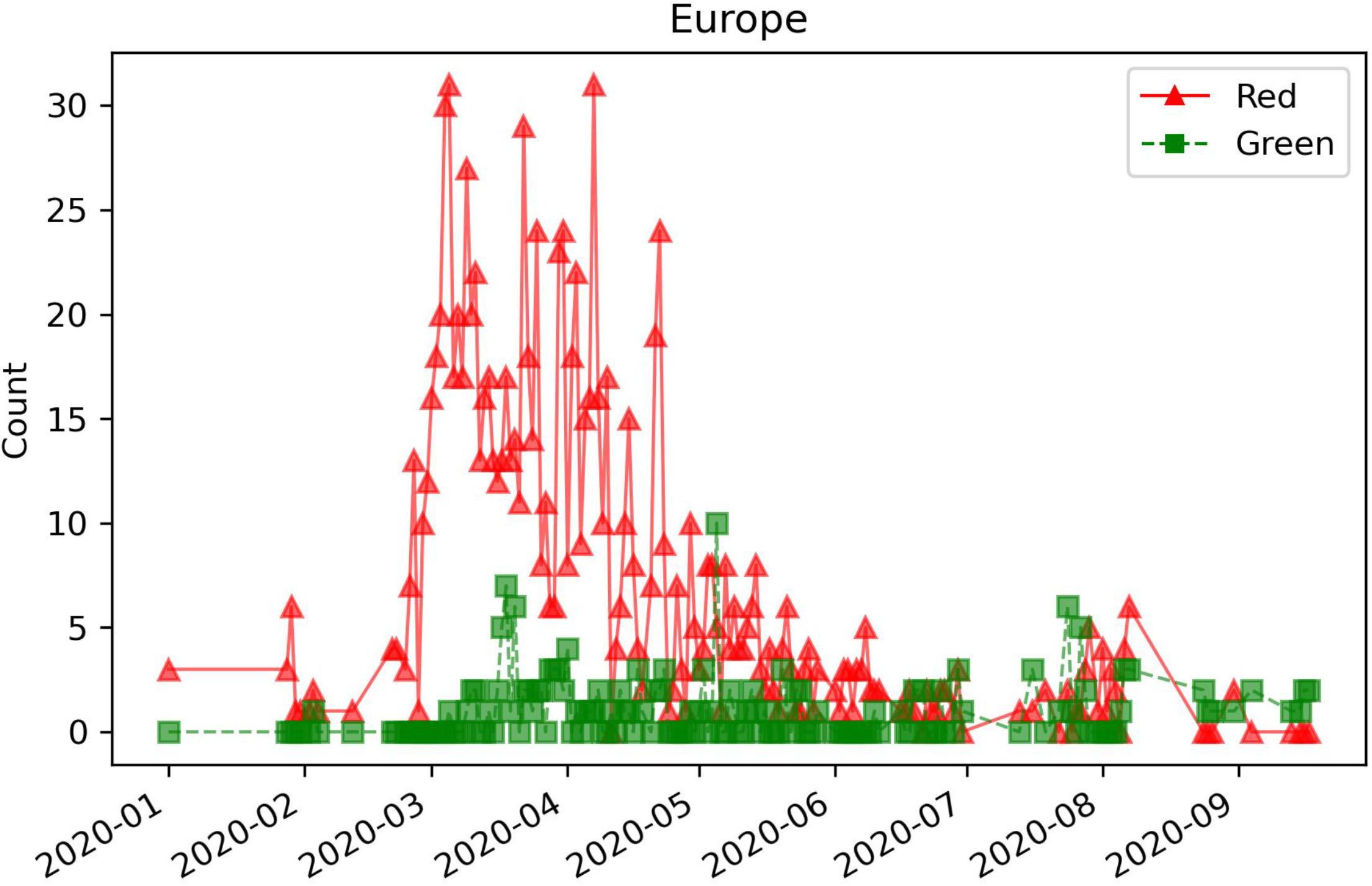
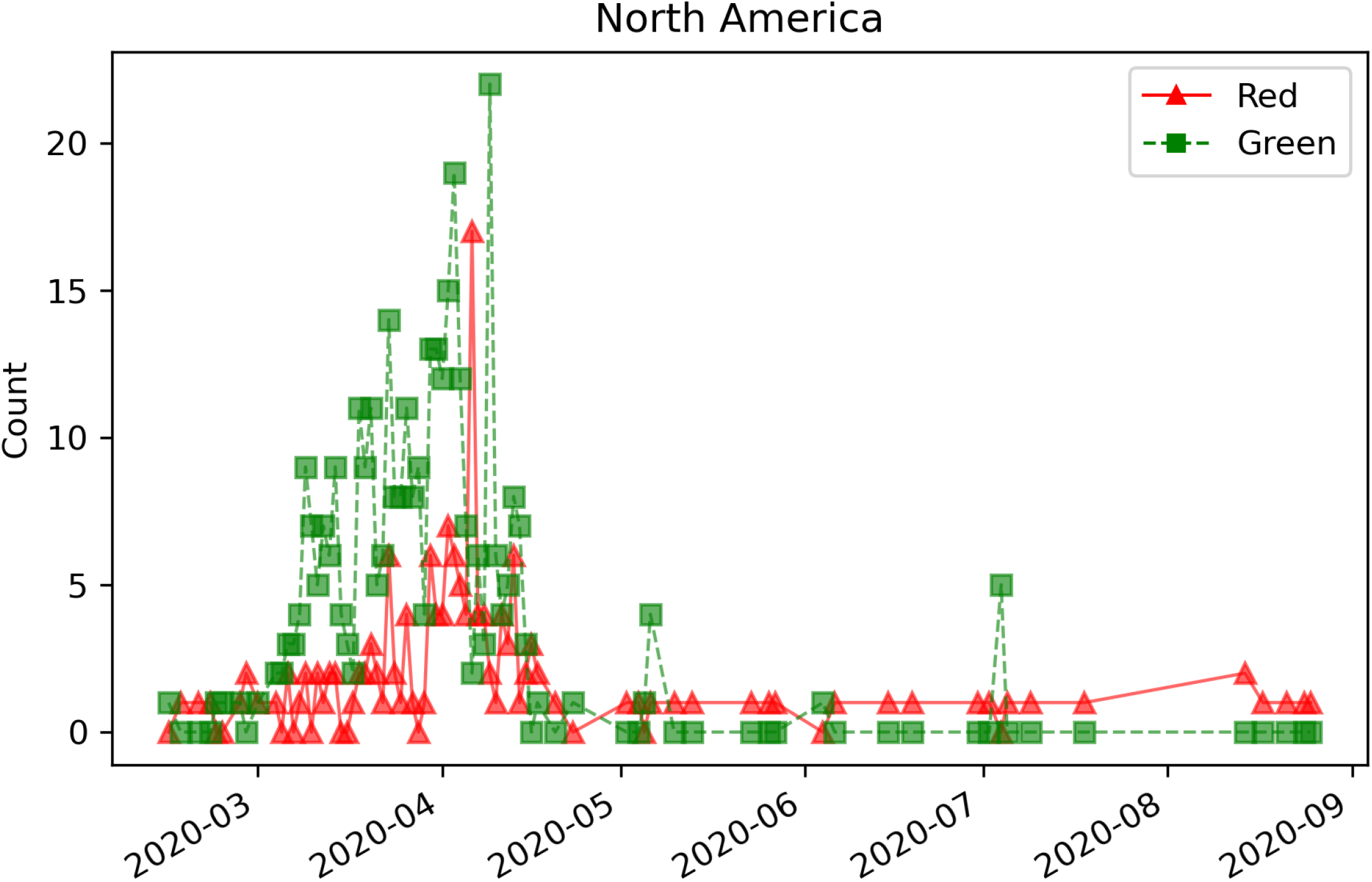
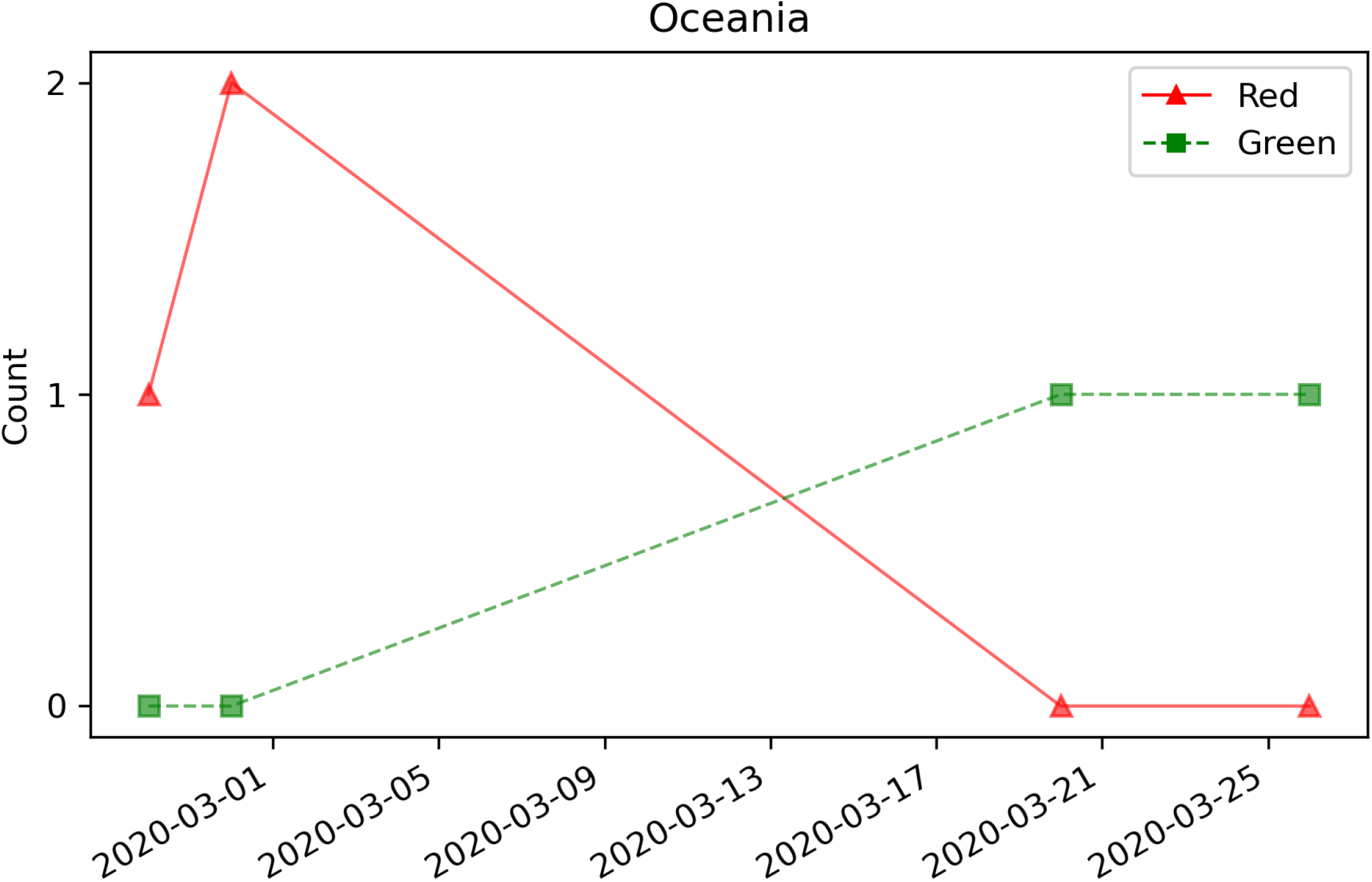
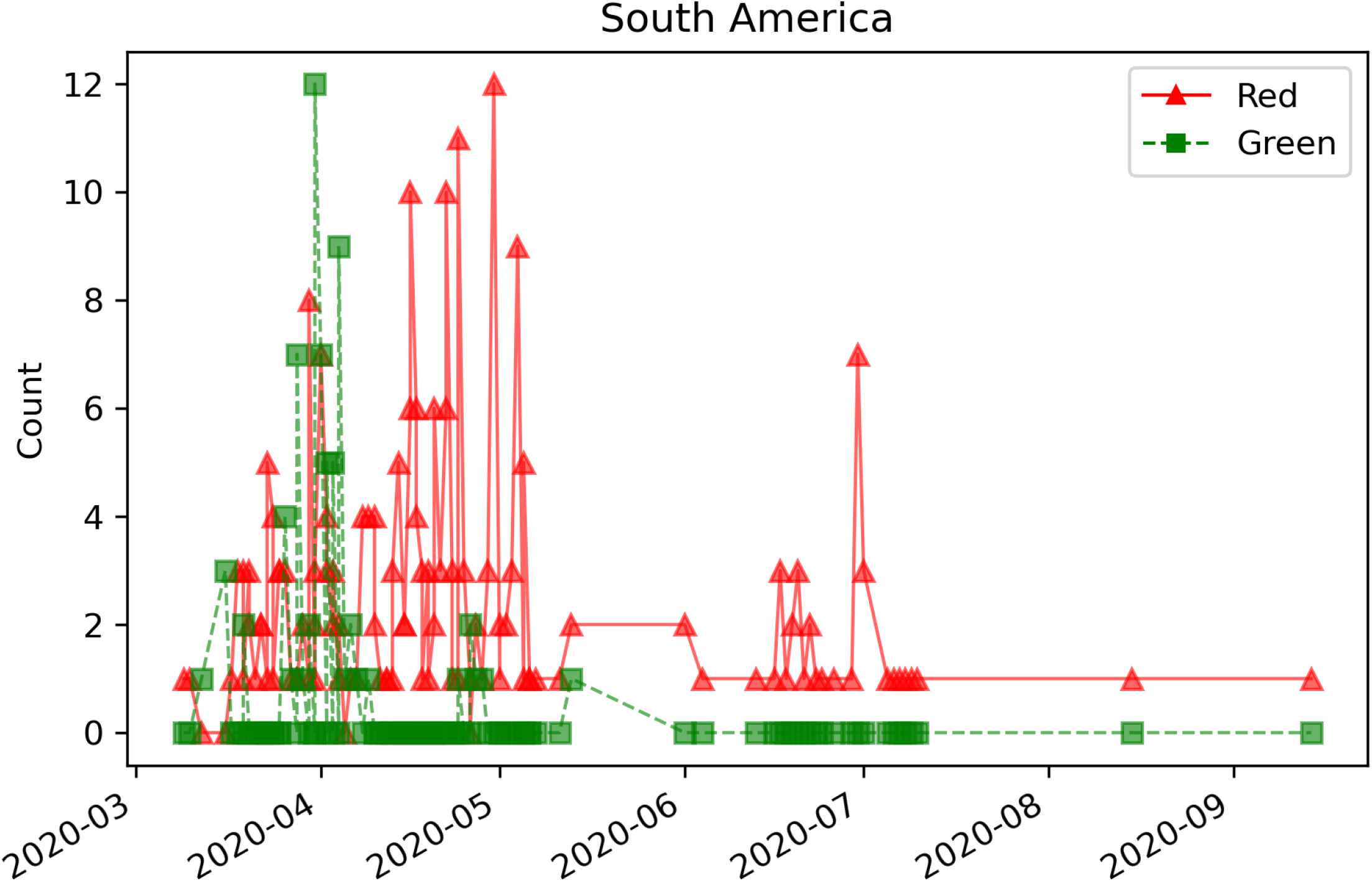
“Severe” and “Mild” outcome counts (n = 3611) over time for all GISAID regions. A) Africa (n = 37) B) Asia (n = 1454) C) Europe (n = 1265) D) North America (n = 499) E) Oceania (n = 5) F) South America (n = 351) Curves are labeled by outcome group (red solid line: “Severe” outcome, green dotted line: “Mild” outcome). Dates (x-axis) are shown in YYYY-MM or YYYY-MM-DD format.

**Figure S3:**
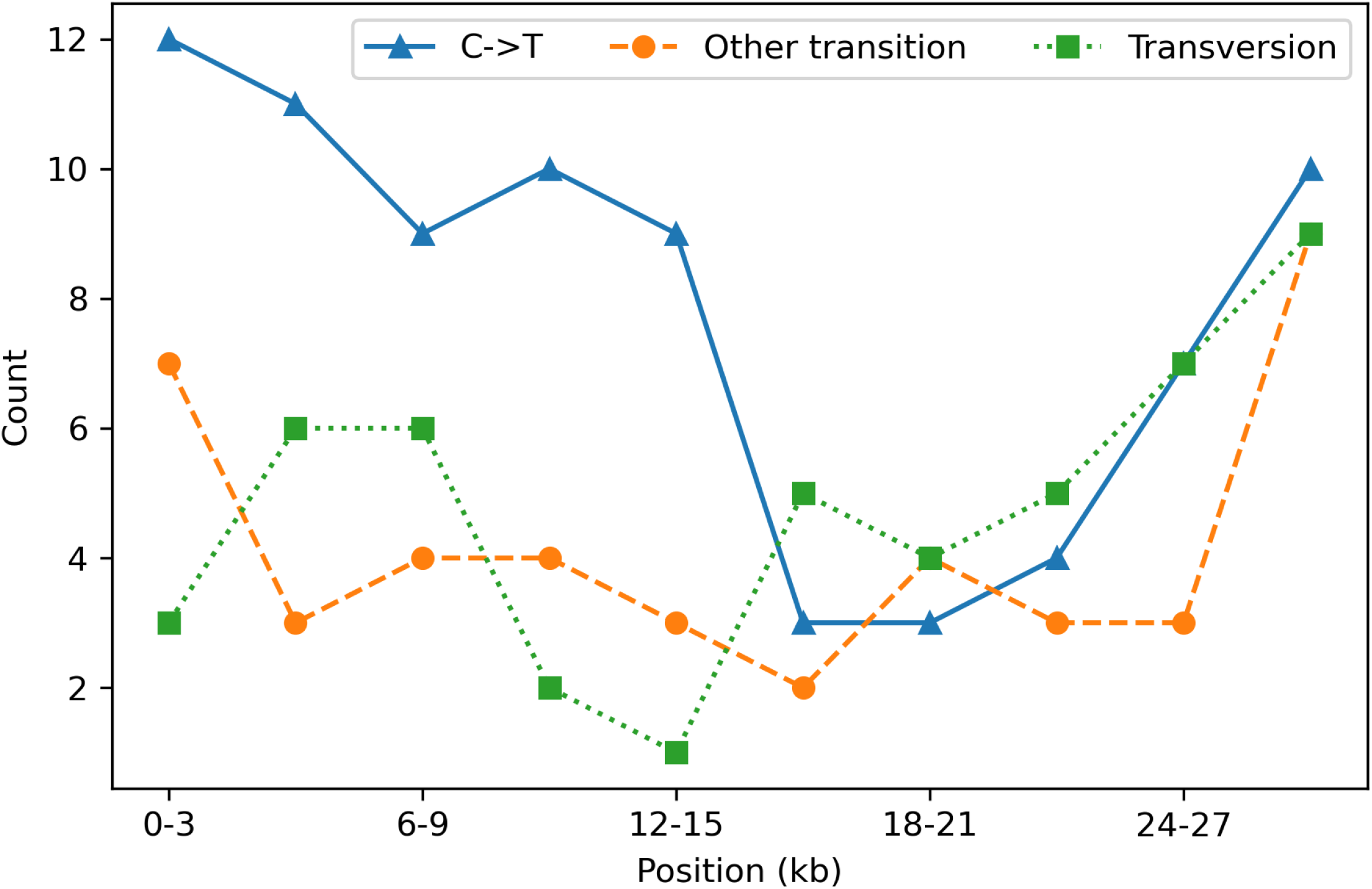
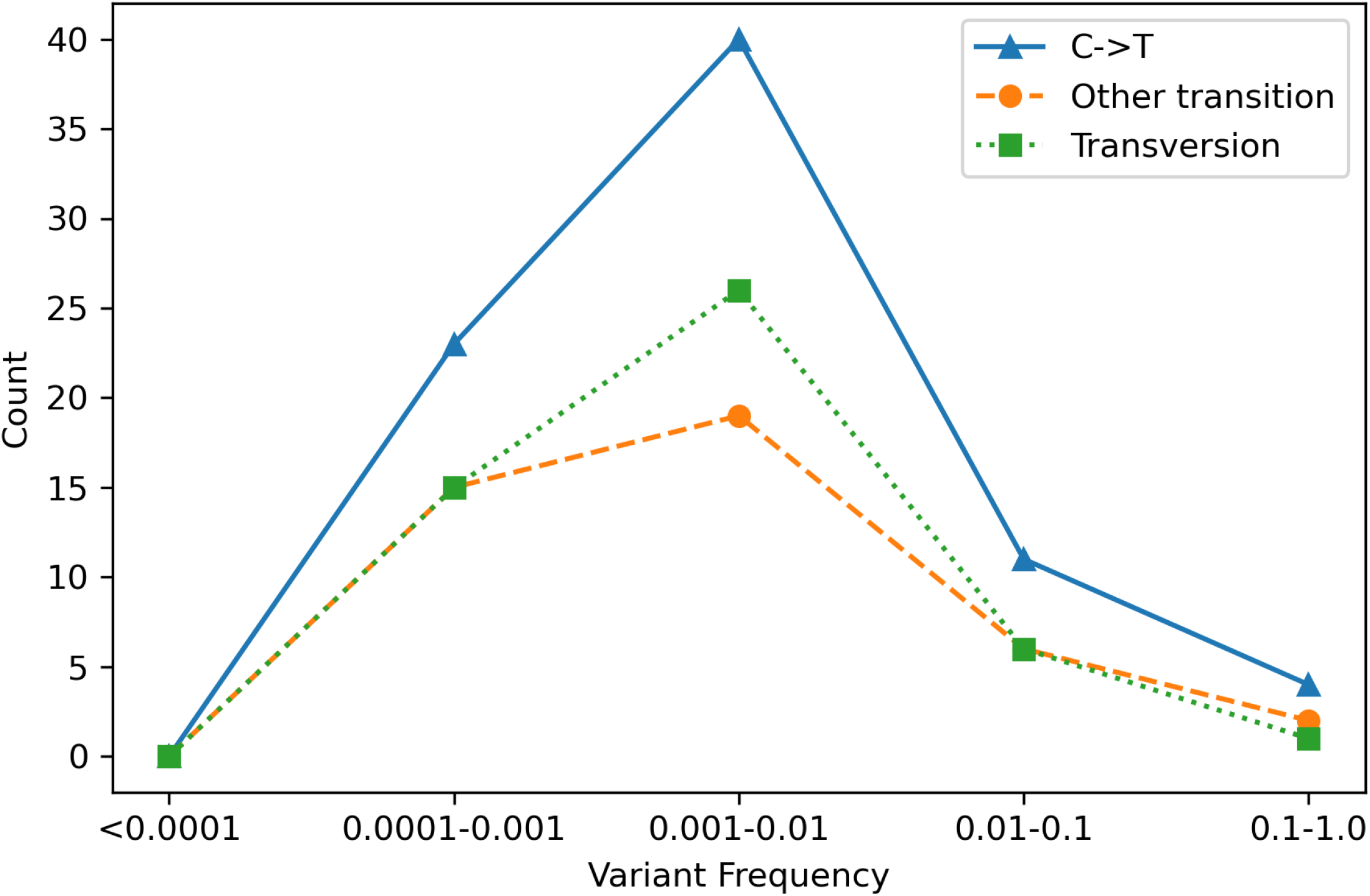
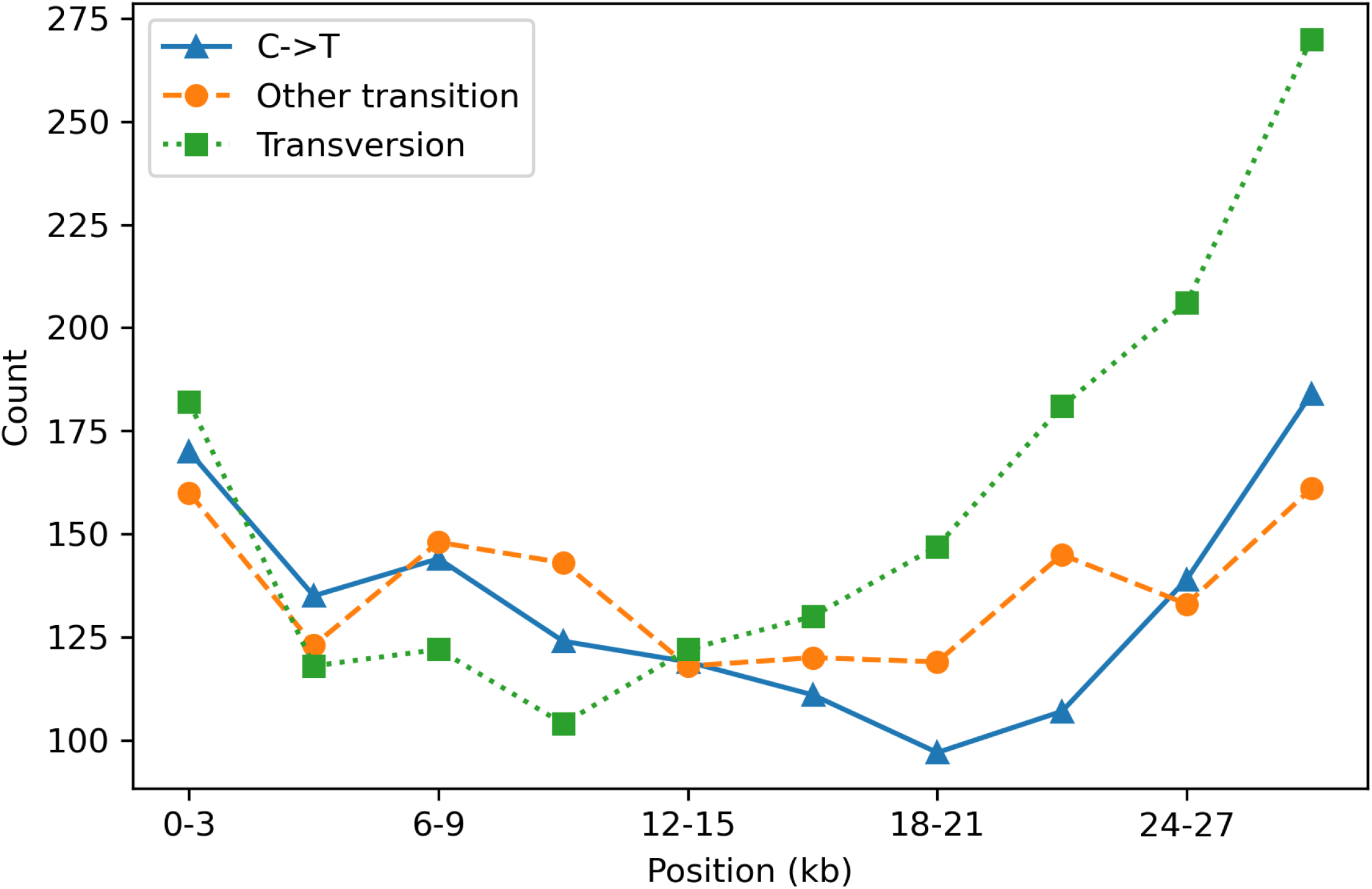
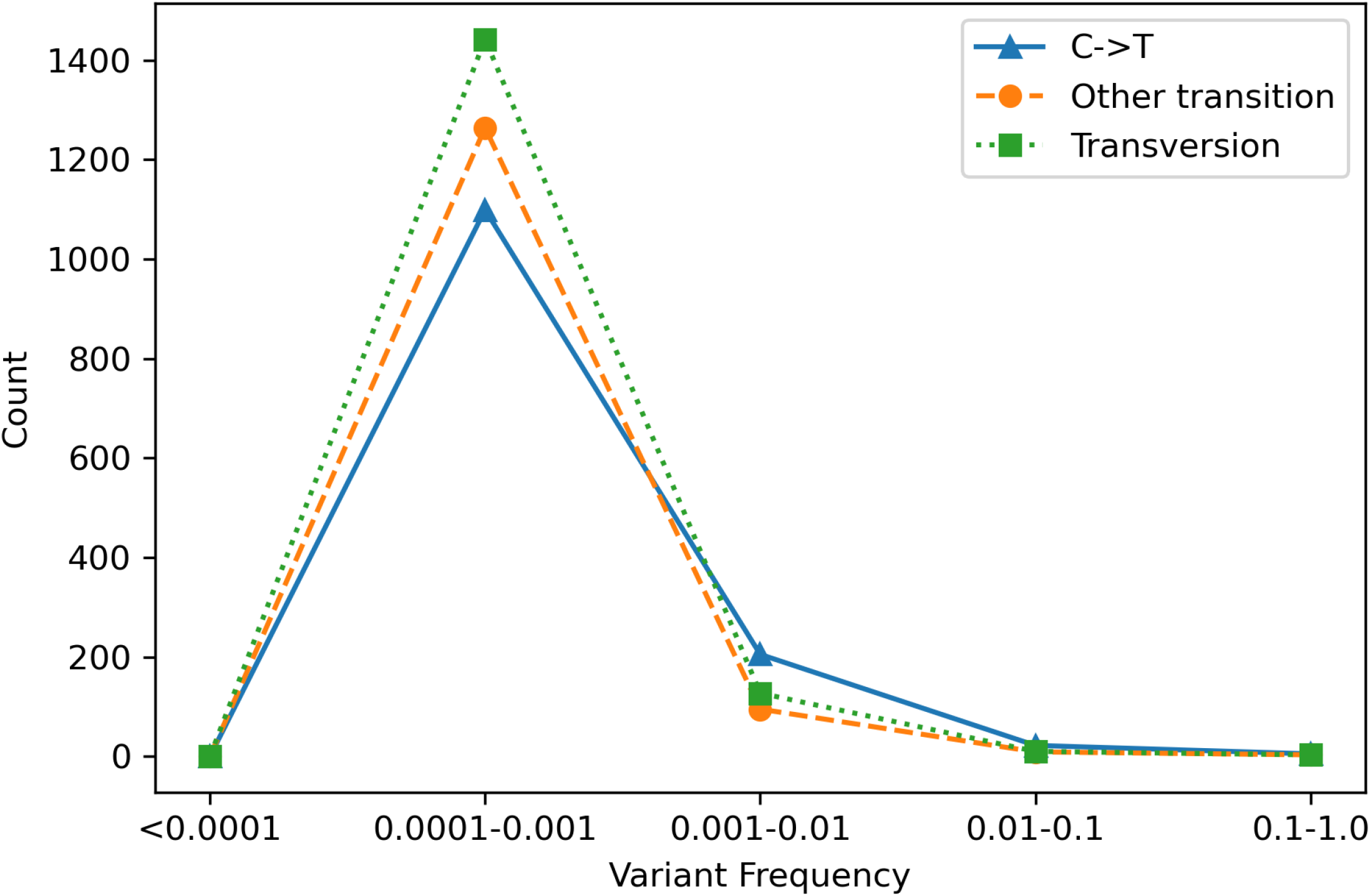
Number of C to T transitions (C->T) compared to other mutation types. A) The counts of variants with odds ratios not equal to one (n=170) plotted against log10 variant frequency half-open intervals, e.g. [0.01, 0.1). B) The counts of variants with odds ratios not equal to one (n=170) plotted against half-open intervals of 3 kilobases (kb), e.g. [0, 3000) in the SARS-CoV-2 genome. C) The counts of variants used for logistic regression modeling (n=4484) plotted against log10 variant frequency half-open intervals, e.g. [0.01, 0.1). D) The counts of variants used for logistic regression modeling (n=4484) plotted against half-open intervals of 3 kilobases (kb), e.g. [0, 3000) in the SARS-CoV-2 genome. Curves are labeled based on mutation type (blue solid line: C->T transition, orange dashed line: other transition [C->T, T->C, A->G, G->A], green dotted line: transversion [C->A, A->C, T->G, G->T, C->G, G->C, A->T, T->A]).

