## Supplementary Table 2 for "Variants in SARS-CoV-2 Associated with Mild or Severe Outcome"

**Supplementary Table 2. Summary of Variants and Association with Severe Outcome.**

| Variant_Name | Odds_Ratio | Amino Acid | Mutation_Type | Variant_Frequency |
|---|---|---|---|---|
| C29625T | 0.01 | S23F | Missense | 0.0015 |
| C5700A | 0.02 | A1812D | Missense | 0.0155 |
| C1281T | 0.02 | A339V | Missense | 0.0039 |
| C10202T | 0.03 | L3313F | Missense | 0.0012 |
| G5554T | 0.04 | K1763N | Missense | 0.0045 |
| C9693T | 0.04 | A3143V | Missense | 0.0015 |
| G26233T | 0.05 | | Non-coding | 0.0018 |
| C12003T | 0.05 | S3913L | Missense | 0.0015 |
| T3833G | 0.06 | F1190V | Missense | 0.0009 |
| G20087C | 0.06 | G6608A | Missense | 0.0012 |
| C2558T | 0.07 | P765S | Missense | 0.0033 |
| C17440T | 0.08 | P5726S | Missense | 0.0021 |
| C14757T | 0.08 | H4831H | Silent | 0.0006 |
| A25596G | 0.09 | R68R | Silent | 0.0012 |
| C8818T | 0.09 | C2851C | Silent | 0.0012 |
| C4993T | 0.09 | N1576N | Silent | 0.0006 |
| C3176T | 0.09 | P971S | Missense | 0.0009 |
| T4092C | 0.09 | I1276T | Missense | 0.0006 |
| G16945A | 0.10 | A5561T | Missense | 0.0015 |
| C3634T | 0.10 | N1123N | Silent | 0.0125 |
| C541T | 0.10 | L92L | Silent | 0.0009 |
| G14181A | 0.10 | L4639L | Silent | 0.0009 |
| C19955T | 0.11 | T6564I | Missense | 0.0006 |
| C10868T | 0.11 | L3535L | Silent | 0.0018 |
| C28311T | 0.14 | P13L | Missense | 0.0999 |
| C16323T | 0.14 | C5353C | Silent | 0.0012 |
| C11563T | 0.14 | C3766C | Silent | 0.0009 |
| G20773T | 0.16 | G6837C | Missense | 0.0006 |
| G28878A | 0.17 | S202N | Missense | 0.0143 |
| A3742G | 0.17 | I1159M | Missense | 0.0015 |
| C1707T | 0.18 | S481F | Missense | 0.0009 |
| T9672A | 0.18 | V3136E | Missense | 0.0006 |
| TGTTA26157T | 0.18 | V256fs | Frameshift | 0.0012 |
| G204T | 0.19 | | Non-coding | 0.0012 |
| G18985T | 0.20 | V6241F | Missense | 0.0009 |
| T16732G | 0.22 | S5490A | Missense | 0.0009 |
| G25429T | 0.23 | V13L | Missense | 0.0033 |
| A26134G | 0.23 | T248A | Missense | 0.0006 |
| T9628C | 0.24 | S3121S | Silent | 0.0006 |
| GCGCTTC26350G | 0.24 | A36_R38delinsG | Deletion | 0.0009 |
| C25466T | 0.24 | P25L | Missense | 0.0021 |
| G28884T | 0.26 | G204V | Missense | 0.0006 |

| | | | | |
|---|---|---|---|---|
| C6354T | 0.27 | S2030L | Missense | 0.0009 |
| C25006T | 0.28 | F1148F | Silent | 0.0015 |
| G29742A | 0.28 | | Non-coding | 0.0161 |
| T8022G | 0.31 | V2586G | Missense | 0.0077 |
| G17808T | 0.32 | K5848N | Missense | 0.0009 |
| C10507T | 0.33 | N3414N | Silent | 0.0054 |
| A6507G | 0.33 | N2081S | Missense | 0.0018 |
| C29774T | 0.35 | | Non-coding | 0.0012 |
| G23236T | 0.36 | K558N | Missense | 0.0021 |
| T23815C | 0.38 | N751N | Silent | 0.0009 |
| T6262C | 0.38 | N1999N | Silent | 0.0006 |
| C13454T | 0.38 | Q4397* | Nonsense | 0.0006 |
| G28321T | 0.39 | T16T | Silent | 0.0045 |
| G29734C | 0.41 | | Non-coding | 0.0059 |
| T23443C | 0.44 | D627D | Silent | 0.0018 |
| C21627T | 0.44 | T22I | Missense | 0.0033 |
| C3927T | 0.46 | S1221L | Missense | 0.0006 |
| T24634C | 0.46 | L1024L | Silent | 0.0006 |
| C25624T | 0.46 | H78Y | Missense | 0.0009 |
| C18877T | 0.47 | L6205L | Silent | 0.1014 |
| C3373A | 0.47 | D1036E | Missense | 0.0039 |
| G25249T | 0.47 | M1229I | Missense | 0.0030 |
| C379A | 0.48 | V38V | Silent | 0.0033 |
| C3874T | 0.50 | I1203I | Silent | 0.0006 |
| C22432T | 0.50 | D290D | Silent | 0.0012 |
| C11620T | 0.51 | F3785F | Silent | 0.0021 |
| C29095T | 0.52 | F274F | Silent | 0.0045 |
| G6894T | 0.53 | C2210F | Missense | 0.0006 |
| G28881A | 0.53 | R203K | Missense | 0.2444 |
| G17014T | 0.54 | D5584Y | Missense | 0.0015 |
| G12167A | 0.55 | V3968I | Missense | 0.0006 |
| G28300T | 0.55 | Q9H | Missense | 0.0006 |
| C12781T | 0.57 | Y4172Y | Silent | 0.0009 |
| A178G | 0.62 | | Non-coding | 0.0006 |
| A28795G | 0.63 | E174E | Silent | 0.0012 |
| G4300T | 0.64 | V1345V | Silent | 0.0056 |
| C12809T | 0.68 | L4182F | Missense | 0.0056 |
| C19018T | 0.70 | P6252S | Missense | 0.0006 |
| C2113T | 0.71 | I616I | Silent | 0.0059 |
| G8653T | 0.73 | M2796I | Missense | 0.0030 |
| A3039G | 0.73 | Y925C | Missense | 0.0012 |
| G21795T | 0.76 | R78M | Missense | 0.0015 |
| A8628G | 0.76 | Y2788C | Missense | 0.0006 |
| A2568G | 0.77 | Q768R | Missense | 0.0006 |
| C27046T | 0.78 | T175M | Missense | 0.0045 |

| | | | | |
|---|---|---|---|---|
| A18052G | 0.79 | T5930A | Missense | 0.0012 |
| G22468T | 0.79 | T302T | Silent | 0.0158 |
| C2416T | 0.80 | Y717Y | Silent | 0.0137 |
| G20134T | 0.83 | V6624L | Missense | 0.0009 |
| T17247C | 0.86 | R5661R | Silent | 0.0039 |
| G2879A | 0.87 | A872T | Missense | 0.0006 |
| C313T | 0.87 | L16L | Silent | 0.0297 |
| C12242T | 0.87 | R3993C | Missense | 0.0009 |
| G3892T | 0.88 | E1209D | Missense | 0.0006 |
| G11083T | 0.88 | L3606F | Missense | 0.1385 |
| C4679T | 0.89 | P1472S | Missense | 0.0009 |
| G8371T | 0.90 | Q2702H | Missense | 0.0042 |
| A19422G | 0.91 | S6386S | Silent | 0.0009 |
| C8352T | 0.93 | A2696V | Missense | 0.0006 |
| G1820A | 0.95 | G519S | Missense | 0.0015 |
| C17403T | 0.96 | A5713A | Silent | 0.0009 |
| C27813T | 0.96 | L20F | Missense | 0.0009 |
| G25641T | 0.96 | L83F | Missense | 0.0006 |
| T312C | 0.97 | L16P | Missense | 0.0006 |
| T27384C | 0.97 | D61D | Silent | 0.0021 |
| T19632C | 0.98 | N6456N | Silent | 0.0006 |
| G15652T | 0.99 | D5130Y | Missense | 0.0018 |
| A17645T | 1.00 | Q5794L | Missense | 0.0009 |
| C6445T | 1.00 | D2060D | Silent | 0.0018 |
| ATG29866A | 1.01 | | Non-coding | 0.0027 |
| A10874G | 1.01 | N3537D | Missense | 0.0018 |
| C26625T | 1.02 | L35L | Silent | 0.0012 |
| C13536T | 1.03 | Y4424Y | Silent | 0.0080 |
| C3317T | 1.05 | P1018S | Missense | 0.0006 |
| G29474T | 1.05 | D401Y | Missense | 0.0015 |
| C1191T | 1.05 | P309L | Missense | 0.0045 |
| G1141T | 1.05 | K292N | Missense | 0.0018 |
| G10360A | 1.05 | K3365K | Silent | 0.0006 |
| T7621C | 1.13 | C2452C | Silent | 0.0065 |
| C875T | 1.15 | L204F | Missense | 0.0012 |
| C4582T | 1.15 | N1439N | Silent | 0.0012 |
| A20268G | 1.17 | L6668L | Silent | 0.0473 |
| C1917T | 1.20 | T551I | Missense | 0.0012 |
| C1059T | 1.21 | T265I | Missense | 0.0847 |
| T13006C | 1.21 | A4247A | Silent | 0.0042 |
| C3037T | 1.23 | F924F | Silent | 0.6153 |
| G28883C | 1.24 | G204R | Missense | 0.2420 |
| A10948G | 1.24 | R3561R | Silent | 0.0024 |
| C10702T | 1.30 | D3479D | Silent | 0.0006 |
| G27415T | 1.34 | A8S | Missense | 0.0027 |

| | | | | |
|---|---|---|---|---|
| C3686T | 1.38 | H1141Y | Missense | 0.0048 |
| G14122T | 1.41 | G4620C | Missense | 0.0027 |
| C23929T | 1.42 | Y789Y | Silent | 0.0990 |
| C28854T | 1.44 | S194L | Missense | 0.0369 |
| C241T | 1.50 | | Non-coding | 0.6201 |
| A29700G | 1.52 | | Non-coding | 0.0036 |
| C14408T | 1.58 | P4715L | Missense | 0.6147 |
| T9172C | 1.58 | L2969L | Silent | 0.0024 |
| C8326T | 1.59 | D2687D | Silent | 0.0042 |
| C27964T | 1.61 | S24L | Missense | 0.0095 |
| C25708T | 1.64 | L106F | Missense | 0.0012 |
| A23403G | 1.71 | D614G | Missense | 0.6118 |
| T28144C | 1.73 | L84S | Missense | 0.0425 |
| G21255C | 1.75 | A6997A | Silent | 0.0012 |
| C8782T | 1.79 | S2839S | Silent | 0.0437 |
| C26750T | 1.80 | I76I | Silent | 0.0065 |
| C9438T | 1.81 | T3058I | Missense | 0.0027 |
| C4543T | 1.82 | T1426T | Silent | 0.0030 |
| G26730T | 1.88 | V70F | Missense | 0.0024 |
| C6040T | 1.93 | F1925F | Silent | 0.0048 |
| C26735T | 2.01 | Y71Y | Silent | 0.0844 |
| T29148C | 2.61 | I292T | Missense | 0.0276 |
| C28076T | 2.62 | C61C | Silent | 0.0009 |
| C28657T | 2.67 | D128D | Silent | 0.0166 |
| G29616T | 2.70 | R20I | Missense | 0.0116 |
| C29870A | 2.73 | | Non-coding | 0.0059 |
| C8139T | 2.92 | S2625F | Missense | 0.0027 |
| C10319T | 3.02 | L3352F | Missense | 0.0042 |
| T27299C | 3.21 | I33T | Missense | 0.0271 |
| C10188T | 3.36 | T3308I | Missense | 0.0033 |
| G29711T | 3.58 | | Non-coding | 0.0009 |
| C23185T | 3.98 | F541F | Silent | 0.0152 |
| C6310A | 4.25 | S2015R | Missense | 0.0488 |
| G26144T | 4.27 | G251V | Missense | 0.0294 |
| C25549T | 5.18 | L53F | Missense | 0.0051 |
| C13620T | 5.86 | D4452D | Silent | 0.0071 |
| G25088T | 74.64 | V1176F | Missense | 0.0369 |
